# Mental Health Concerns Related to the COVID-19 Pandemic on Twitter in the UK

**DOI:** 10.1101/2021.09.27.21264177

**Authors:** Daiwei Zhang, Yue Liu, Senqi Zhang, Li Sun, Pin Li, Ajay Anand, Zidian Xie, Dongmei Li

## Abstract

**Background:** Amid the COVID-19 pandemic, mental health-related symptoms (such as depression and anxiety) have been actively mentioned on social media.

**Objective:** In this study, we aimed to monitor mental health concerns on Twitter during the COVID-19 pandemic in the United Kingdom (UK), and assess the potential impact of the COVID-19 pandemic on mental health concerns of Twitter users.

**Methods:** We collected COVID-19 and mental health-related tweets from the UK between March 5, 2020 and January 31, 2021 through the Twitter Streaming API. We conducted topic modeling using Latent Dirichlet Allocation model to examine discussions about mental health concerns. Deep learning algorithms including Face++ were used to infer the demographic characteristics (age and gender) of Twitter users who expressed mental health concerns related to the COVID-19 pandemic.

**Results:** We showed a positive correlation between COVID-19-related mental health concerns on Twitter and the severity of the COVID-19 pandemic in the UK. Geographic analysis showed that populated urban areas have a higher proportion of Twitter users with mental health concerns compared to England as a whole. Topic modeling showed that general concerns, COVID-19 skeptics, and Death toll were the top topics discussed in mental health-related tweets. Demographic analysis showed that middle-aged and older adults might be more likely to suffer from mental health issues or express their mental health concerns on Twitter during the COVID-19 pandemic.

**Conclusions:** The COVID-19 pandemic has noticeable effects on mental health concerns on Twitter in the UK, which varied among demographic and geographic groups.

## Introduction

On March 11, 2020, the World Health Organization officially announced the COVID-19, the contagious disease caused by the novel coronavirus SARS-CoV-2, as a pandemic. In the United Kingdom, for example, there were over 4.3 million confirmed COVID-19 cases and more than 120,000 associated deaths reported up to the March of 2021 ever since the first case was confirmed in York in England on January 22, 2020 ^1^. The COVID-19 pandemic is widely disruptive to the whole society, especially the economy ^2^. For example, United Kingdom has gone through three lockdowns, and many businesses have been temporarily or permanently shut down. Even worse, both the pandemic itself and the economic disruption had a serious impact on people’s mental health. It was estimated in 2014 that one in six adults in England had a common mental disorder, and mental disorders were more common in people living alone ^3^. Due to the factors like isolations and restrictions, economic recession, or even the neurological and psychiatric sequelae of COVID-19 ^4^, mental health in the United Kingdom had significantly deteriorated during the early stage of the pandemic compared to the pre-COVID-19 era ^5^. While recent studies showed that the evolving COVID-19 pandemic leads to mental health globally with increasing cases of depression, self-harm, suicide, etc. ^6 7^, it is crucial to identify the individuals at risk of mental health issues as early as possible so that substantial resources could be better allocated.

Twitter, one of the most widely used social networking services globally, provides a rich data source for studying mental health during the COVID-19 pandemic. A pre-pandemic study has shown that the application of natural language processing techniques on Twitter data could provide insights into several specific mental health disorders ^8^. Since the outbreak of COVID-19 led to a burst of information in social media, many studies, mostly within the United States, have been conducted to monitor and analyze the tweets (i.e., Twitter posts) with a specific focus on people’s mental health during the pandemic ^9 10 11^.

In this study, we specifically focused on the mental health-related tweets associated with the COVID-19 pandemic in the United Kingdom. We aimed to understand potential mental health concerns expressed on Twitter during the first two waves of the UK’s pandemic temporally and geographically. Furthermore, we employed demographic inference to characterize Twitter users with mental health concerns.

## Methods

### COVID-19 Pandemic Data Collection

We obtained the daily new COVID-19 cases from March 4, 2020 to January 31, 2021 from the COVID-19 datasets published and updated by GOV.UK [https://coronavirus.data.gov.uk], as well as the United Kingdom public sector information website.

### COVID-19 Twitter Data Collection and Preprocessing

We collected COVID-19-related tweets posted between March 5, 2020 and January 31, 2021 through the Twitter Streaming API using the keywords (“corona”, “covid19”, “covid”, “coronavirus”, and “ncov”) ^12^. The Twitter data from May 18 to May 20, 2020, and from August 25 to September 14, 2020 were missing due to some technical failure. Duplicate tweets and retweets were removed. As “corona” is one of the top-selling beer brands, we removed the promotional and commercial posts, which mentioned words “dealer,” “supply,” “beer,” “discount,” “sale,” etc. After data cleaning, in order to obtain mental health-related tweets, we further filtered the COVID-19-related tweets with a list of 27 keywords, including “depression”, “depressed”, “depress”, “failure”, “hopeless”, “nervous”, “restless”, “tired”, “worthless”, “unrested”, “fatigue”, “irritable”, “stress”, “dysthymia”, “anxiety”, “adhd”, “loneliness”, “lonely”, “alone”, “boredom”, “boring”, “fear”, “worry”, “anger”, “confusion”, “insomnia”, and “distress” ^13^. A total of 5,088,049 COVID-19 and mental health-related tweets in English were obtained.

To identify the tweets from the United Kingdom based on their tagged geolocation, we constructed a comprehensive list of the UK’s geographic divisions with the names of four constituent countries (England, Scotland, Wales, and Northern Ireland) and their administrative subdivisions, which includes regions (e.g. East Midlands as one of nine official regions of England), counties (e.g. Greater Manchester as the metropolitan county), districts (e.g. Birmingham as the Metropolitan district), boroughs (e.g. London borough), and cities (e.g. London). Considering many places in the United States were named after places in the UK, such as Manchester, Birmingham, and Rochester, we further removed the tweets with geolocation that contains “American” characteristics, including the terms “United States”, “US”, the names of states, and the capitalized state abbreviations (e.g., NY and CA). Finally, a dataset of 317,292 mental health-related tweets posted from 148,534 unique Twitter users in the United Kingdom, was obtained for our further analysis.

We assigned the mental health-related tweets to subsets of four countries (England, Scotland, Wales, and Northern Ireland) and three most populated metropolitan areas (London, Birmingham, and Manchester) based on the geolocation information in each tweet.

### Topic Modeling

To identify the most frequent topics discussed in our dataset, we applied the unsupervised Latent Dirichlet allocation (LDA) model ^14^, which is a generative probabilistic model widely used for collections of text corpora. The LDA model can allocate every word in the corpus to a specific topic, where each word in the topic is given a weight value based on its appearance.

Before applying the LDA model for text preprocessing, we first removed the noise, including email addresses, URLs, etc., and tokenized the text by removing all punctuations and converting sentences into a list of lowercase tokens. Then, we removed the stop words defined by Natural Language Toolkit (NLTK) library, including “a”, “the”, etc. Bigrams were then identified, and words were lemmatized to their base forms by the spaCy library. Finally, we constructed the corpus and dictionary on the preprocessed dataset and input them into the LDA algorithm implemented in the Genism package. Models with different parameters of topic numbers were built, and the topic coherence score of each model was used to determine the optimal number of latent topics in our dataset.

### Demographics Inference

From the 148,534 UK Twitter users with mental health concerns, we identified 39,294 valid profile images with Face++ Detect API ^15^. Specifically, a Twitter user profile image is considered to be valid if its URL is accessible and the image contains exactly one recognizable human face. We estimated the gender and age from the attributes returned by Detect API for each valid profile image.

## Results

### Temporal Analysis of Mental Health Concerns on Twitter in the UK

To understand the dynamic change in mental health concerns on Twitter in the UK during the COVID-19 pandemic, we examined a daily number of mental health-related tweets, as well as daily new COVID-19 cases from March 5, 2020 to January 31, 2021. As shown in Figure 1, there were three major waves of COVID-19 cases in the United Kingdom: the first wave was between March and May in 2020; the second one was between late August and November in 2020; the third one was between early December in 2020 and January in 2021. Meanwhile, we observed that there were three major waves of mental health-related tweets, which show similar patterns as the daily COVID-19 cases.

**Figure 1.**
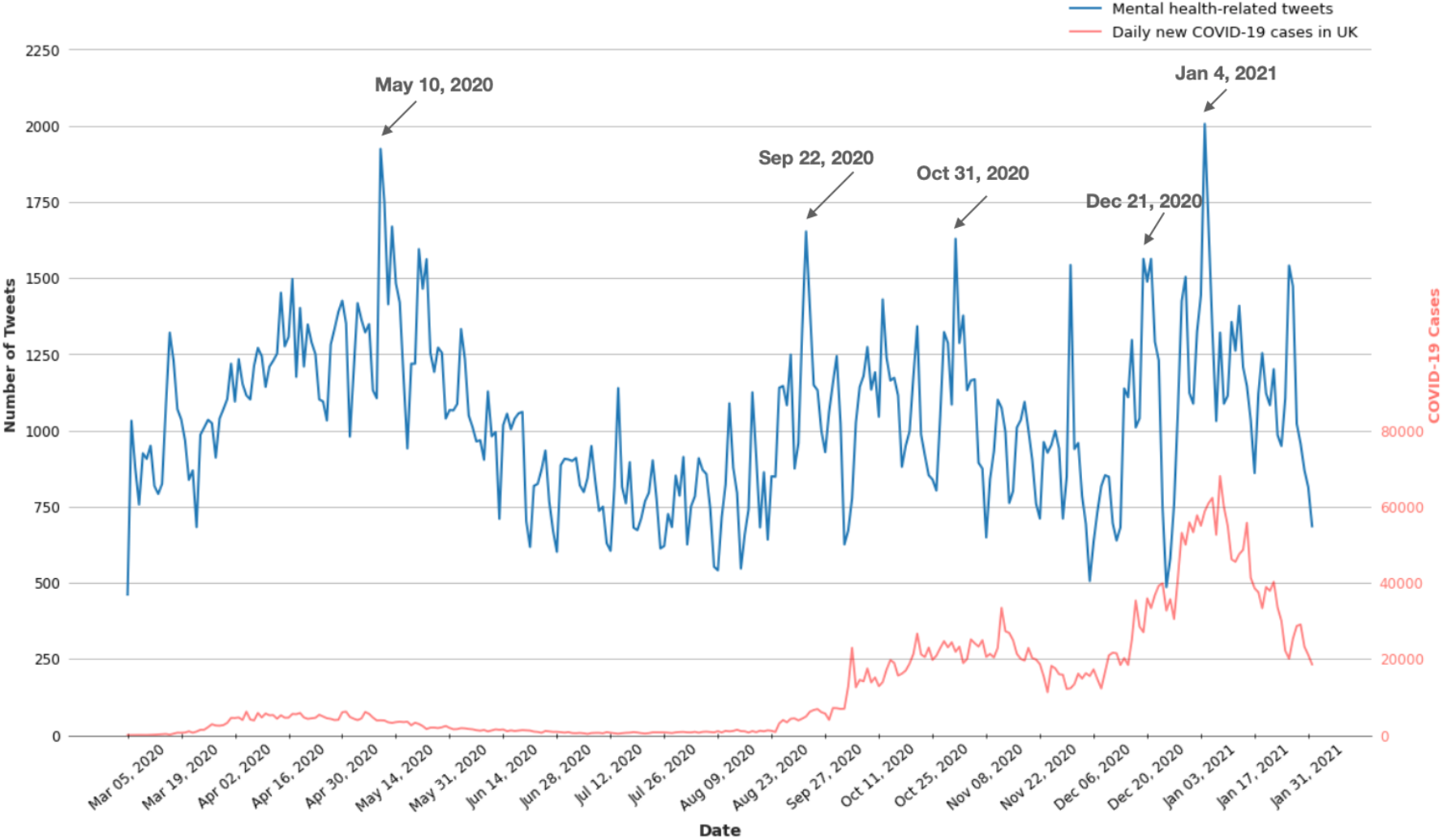
Daily number of mental health-related tweets and COVID-19 new cases in the UK.

The grey arrows in Figure 1 indicate five peaks (including May 10, 2020, September 22, 2020, October 31, 2020, December 21, 2020, and January 4, 2021) with a relatively high number of mental health-related tweets. We further examined the archived UK news released within the 24-hour period for these five peaks. Most of the news is about government responses to control the COVID-19 pandemic, particularly the announcements of restrictions or lockdowns (Supplemental Table 1).

### Geographic Distribution of Twitter Users with Mental Health Concerns in the UK

To understand whether there were some geographic differences in mental health concerns in the UK, we calculated the number of distinct Twitter users who had posted mental health-related tweets by regions or urban areas and normalized them to the population. As shown in Figure 2, we observed that Scotland has the highest proportion of Twitter users with mental health concerns among four countries in the UK. In addition, populated urban areas have a higher proportion of Twitter users with mental health concerns. For example, there were 4,193 Twitter users/million people with mental health concerns in London compared to 1,730 Twitter users/million people in England as a whole (Figure 2).

**Figure 2.**
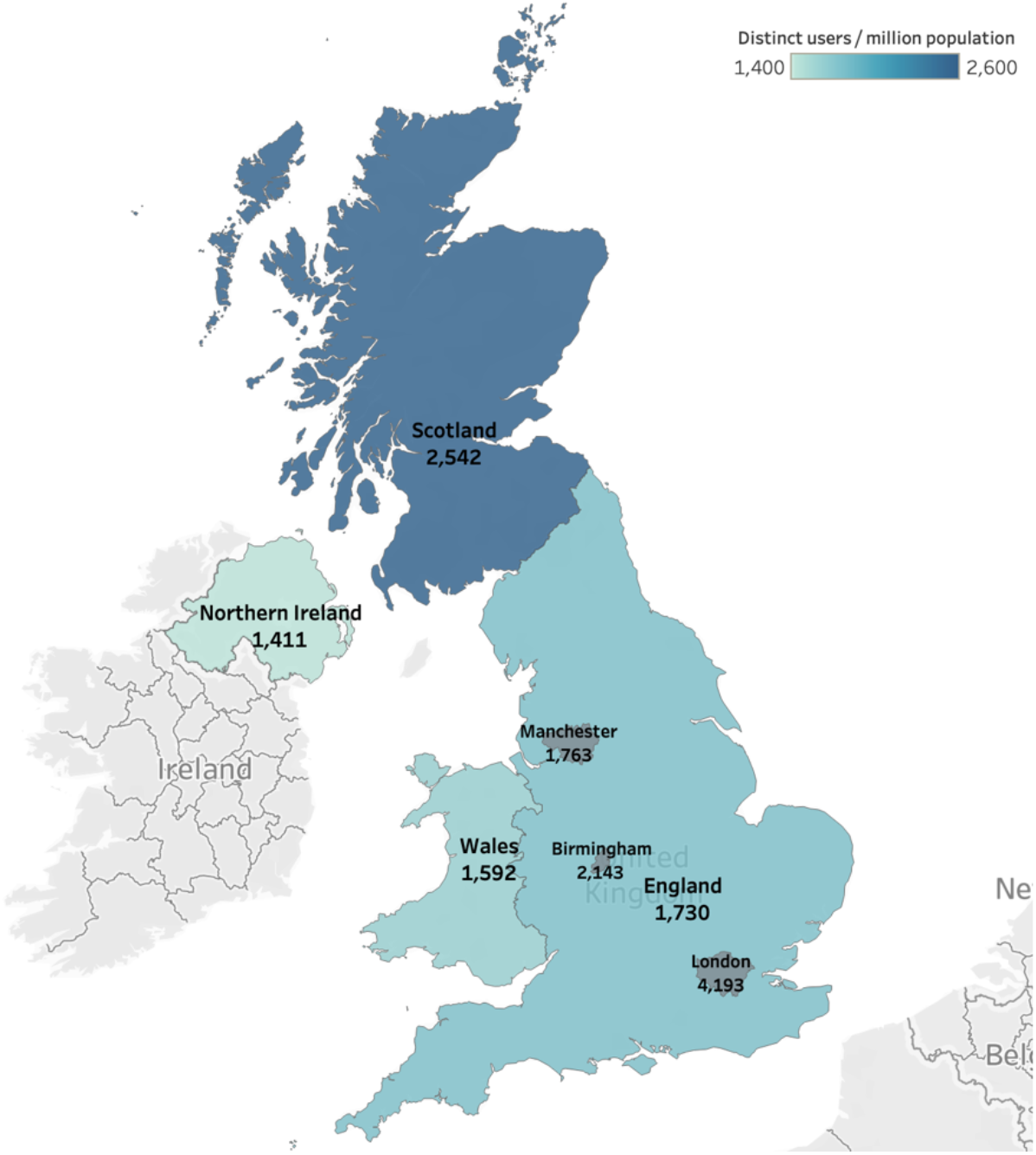
Geographic differences in Twitter users who had mental health concerns in the UK.

### Major Topics Discussed in Mental Health-Related Tweets

To obtain content-wise insights into what topics of discussion are often associated with mental health issues, the LDA model was trained on our tweet dataset for topic modeling. Table 1 showed the major 10 topics identified by our model. As shown, the general concern about pandemic for its uncertainty and the impact on daily life is the most frequently discussed topic (14.4%), followed by the coronavirus skeptics (12.7%) that argue that coronavirus isn’t a real concern but a government tool to spread fear. It’s noteworthy that the topic regarding death toll (10.9%) includes two opposing points of view: the death toll is overwhelming and depressing while the other view thinks the number of deaths is significantly exaggerated by the authority.

**Table 1.**
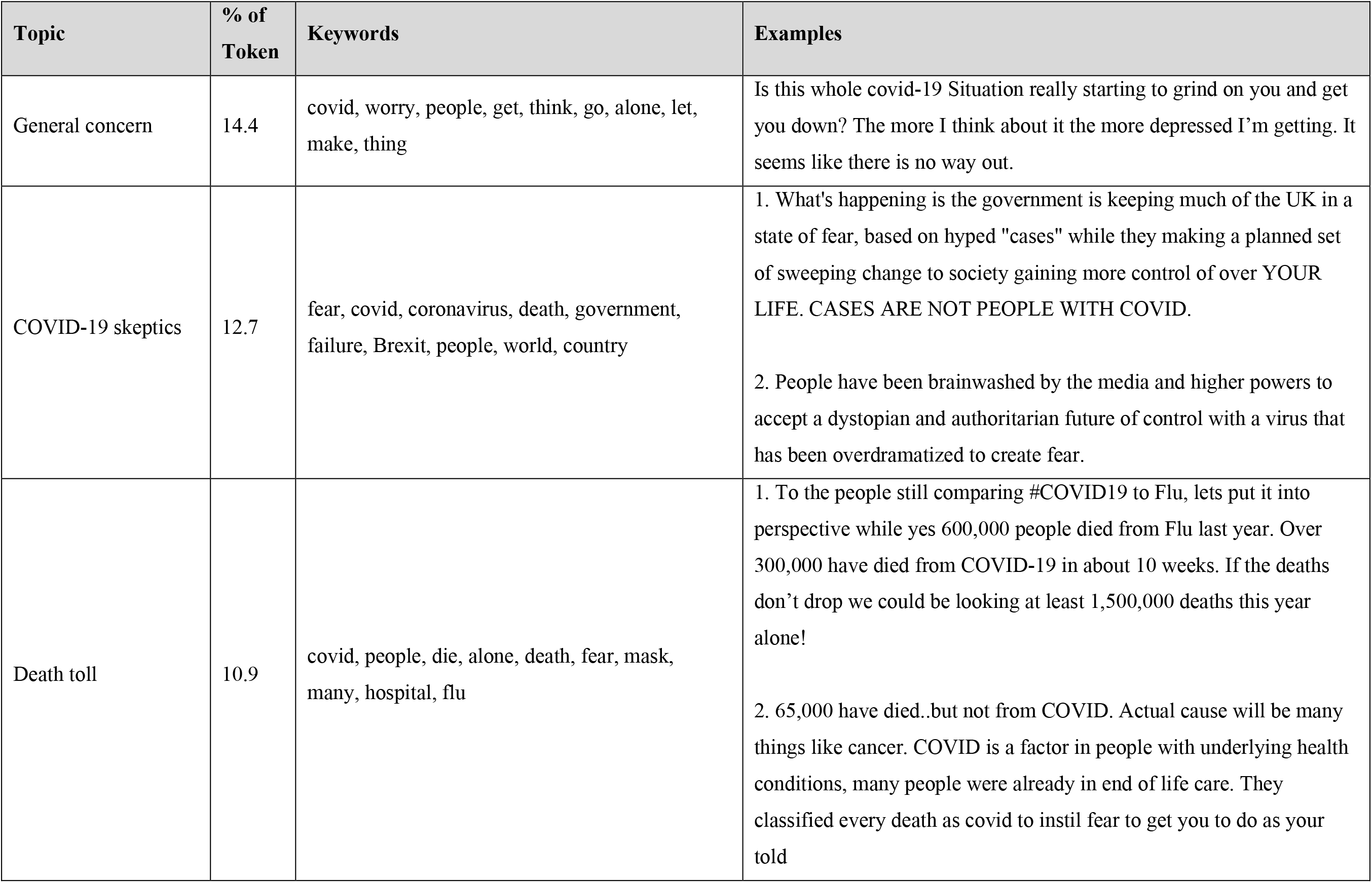
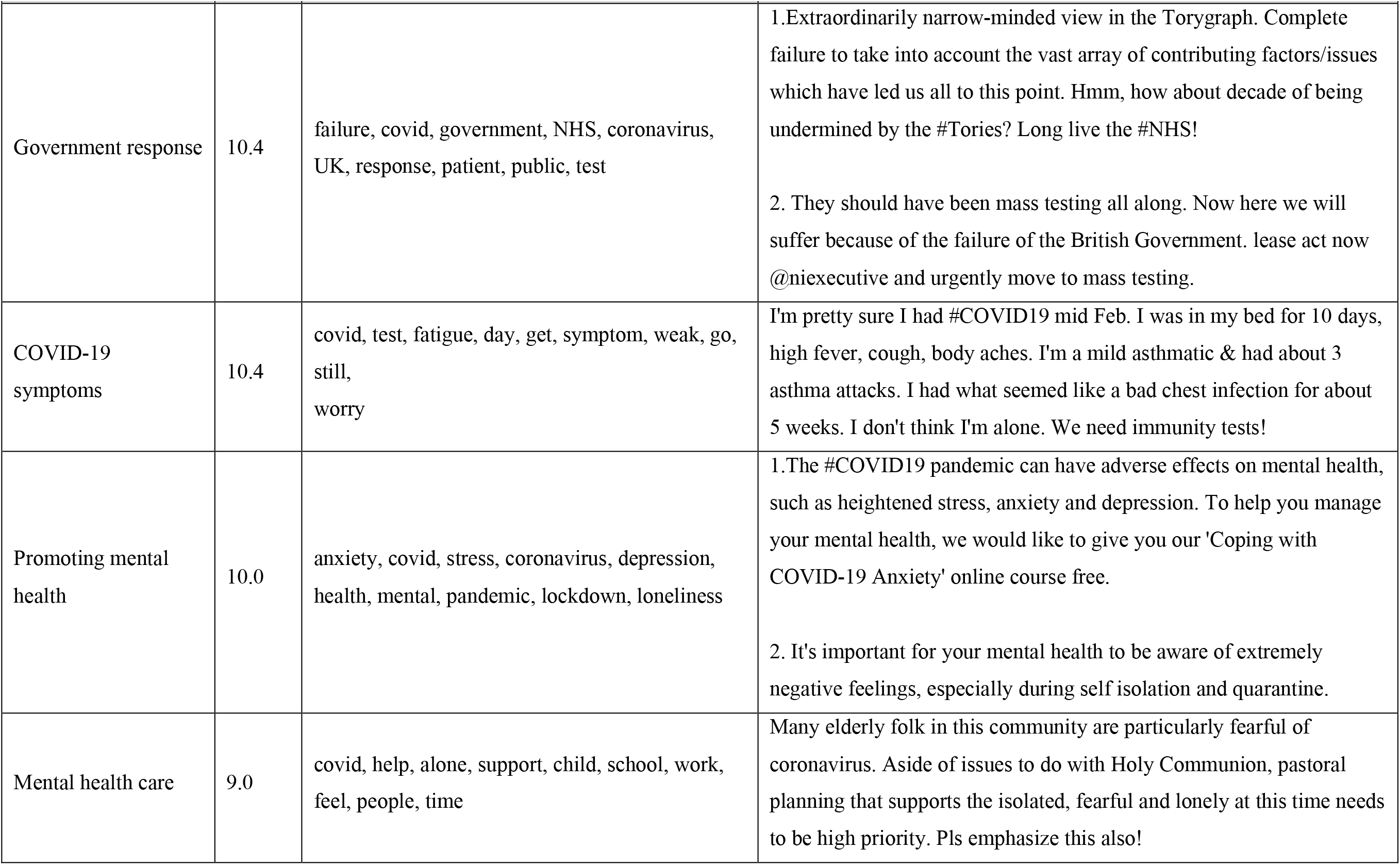
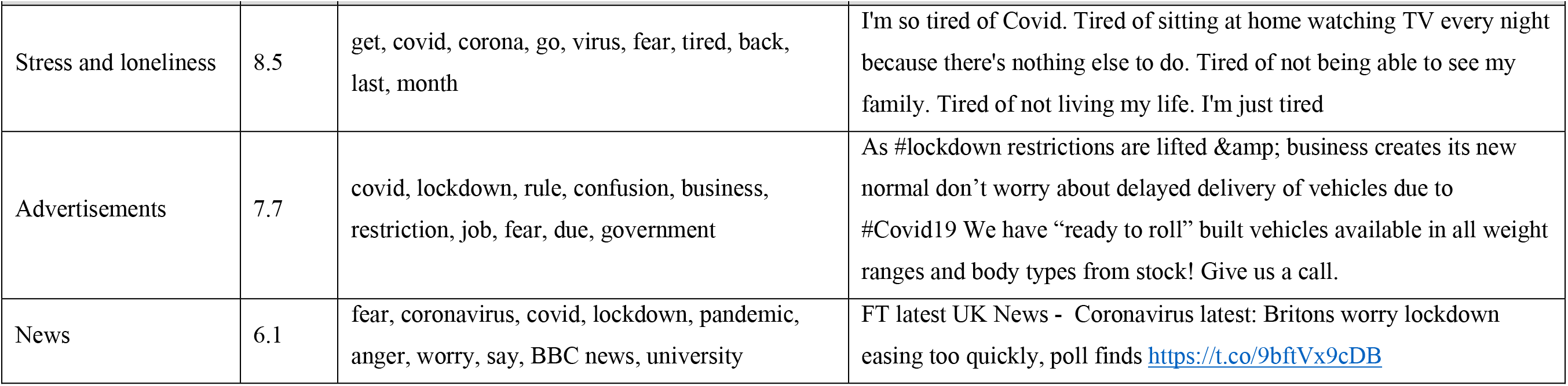
Major topics discussed in COVID-19 and mental health-related tweets from the United Kingdom

| Topic | % of Token | Keywords | Examples |
| --- | --- | --- | --- |
| General concern | 14.4 | covid, worry, people, get, think, go, alone, let, make, thing | Is this whole covid-19 Situation really starting to grind on you and get you down? The more I think about it the more depressed I'm getting. It seems like there is no way out. |
| COVID-19 skeptics | 12.7 | fear, covid, coronavirus, death, government, failure, Brexit, people, world, country | 1. What's happening is the government is keeping much of the UK in a state of fear, based on hyped "cases" while they making a planned set of sweeping change to society gaining more control of over YOUR LIFE. CASES ARE NOT PEOPLE WITH COVID.<br>2. People have been brainwashed by the media and higher powers to accept a dystopian and authoritarian future of control with a virus that has been overdramatized to create fear. |
| Death toll | 10.9 | covid, people, die, alone, death, fear, mask, many, hospital, flu | 1. To the people still comparing #COVID19 to Flu, lets put it into perspective while yes 600,000 people died from Flu last year. Over 300,000 have died from COVID-19 in about 10 weeks. If the deaths don't drop we could be looking at least 1,500,000 deaths this year alone!<br>2. 65,000 have died..but not from COVID. Actual cause will be many things like cancer. COVID is a factor in people with underlying health conditions, many people were already in end of life care. They classified every death as covid to instil fear to get you to do as your told |
| Government response | 10.4 | failure, covid, government, NHS, coronavirus, UK, response, patient, public, test | 1.Extraordinarily narrow-minded view in the Torygraph. Complete failure to take into account the vast array of contributing factors/issues which have led us all to this point. Hmm, how about decade of being undermined by the #Tories? Long live the #NHS!<br>2. They should have been mass testing all along. Now here we will suffer because of the failure of the British Government. lease act now @niexecutive and urgently move to mass testing. |
| COVID-19 symptoms | 10.4 | covid, test, fatigue, day, get, symptom, weak, go, still, worry | I'm pretty sure I had #COVID19 mid Feb. I was in my bed for 10 days, high fever, cough, body aches. I'm a mild asthmatic & had about 3 asthma attacks. I had what seemed like a bad chest infection for about 5 weeks. I don't think I'm alone. We need immunity tests! |
| Promoting mental health | 10.0 | anxiety, covid, stress, coronavirus, depression, health, mental, pandemic, lockdown, loneliness | 1.The #COVID19 pandemic can have adverse effects on mental health, such as heightened stress, anxiety and depression. To help you manage your mental health, we would like to give you our 'Coping with COVID-19 Anxiety' online course free.<br>2. It's important for your mental health to be aware of extremely negative feelings, especially during self isolation and quarantine. |
| Mental health care | 9.0 | covid, help, alone, support, child, school, work, feel, people, time | Many elderly folk in this community are particularly fearful of coronavirus. Aside of issues to do with Holy Communion, pastoral planning that supports the isolated, fearful and lonely at this time needs to be high priority. Pls emphasize this also! |
| Stress and loneliness | 8.5 | get, covid, corona, go, virus, fear, tired, back, last, month | I'm so tired of Covid. Tired of sitting at home watching TV every night because there's nothing else to do. Tired of not being able to see my family. Tired of not living my life. I'm just tired |
| Advertisements | 7.7 | covid, lockdown, rule, confusion, business, restriction, job, fear, due, government | As #lockdown restrictions are lifted & business creates its new normal don't worry about delayed delivery of vehicles due to #Covid19 We have "ready to roll" built vehicles available in all weight ranges and body types from stock! Give us a call. |
| News | 6.1 | fear, coronavirus, covid, lockdown, pandemic, anger, worry, say, BBC news, university | FT latest UK News - Coronavirus latest: Britons worry lockdown easing too quickly, poll finds https://t.co/9bftVx9cDB |

### Demographic Characteristics of Twitter Users with Mental Health Concerns

To understand which demographic groups might be more likely to have mental health concerns on Twitter, we estimated the demographics of Twitter users with mental health concerns. Among Twitter users who had mental health concerns in the UK, 56.86% are male, and 43.14% are female (Figure 3). In addition, Twitter users with age 21-30 were the most (25.03%) among Twitter users who had mental health concerns, followed by the age 31-40 group (23.22%) and age 41-50 group (20.25%). The age 13-20 group was the least one (3.03%).

**Figure 3.**
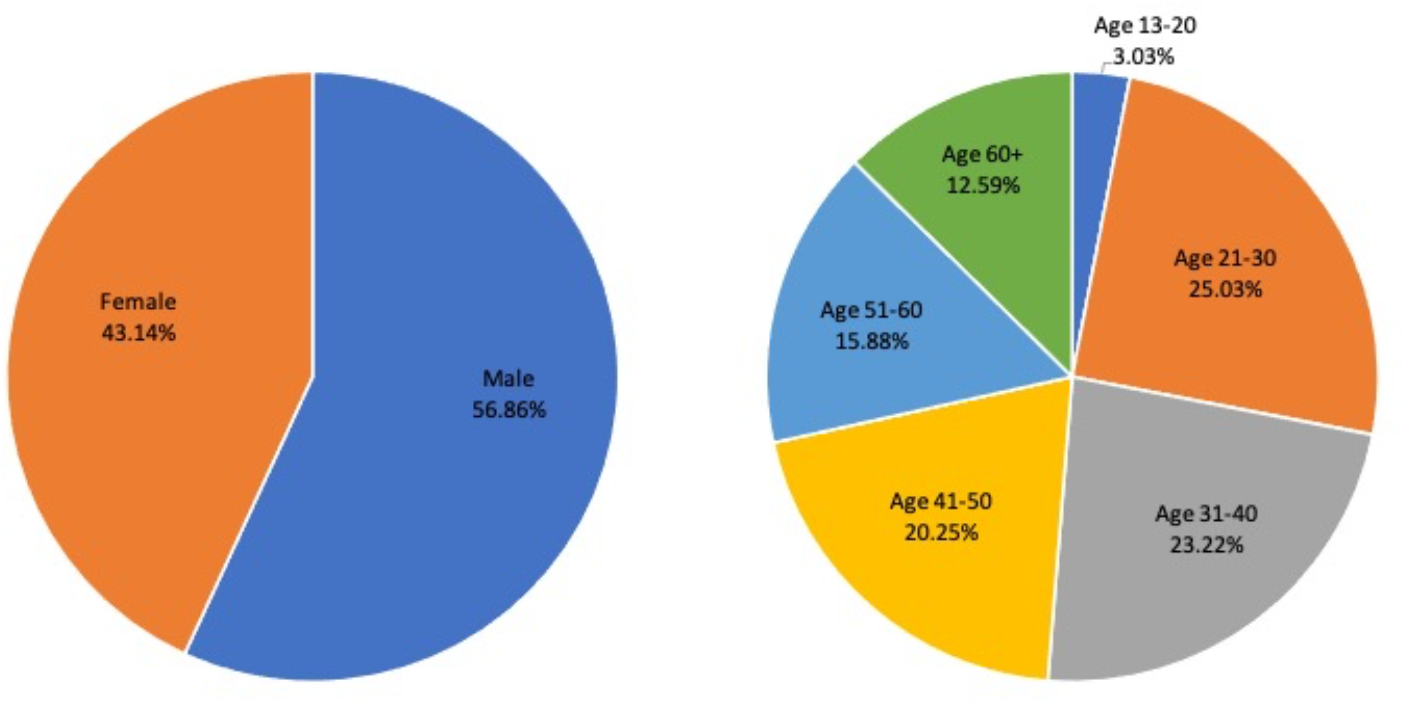
Age and gender composition for Twitter Users with mental health concerns

## Discussion

Through the temporal analysis on COVID-19 and mental health-related tweets in the UK, we observed that there were three major waves in mental health discussion on Twitter, which coincided with three waves of the COVID-19 pandemic in the UK. Furthermore, we showed that the announcements of restrictions measures to control the pandemic by the prime minister, especially the lockdowns, often led to a peak in the number of mental health-related tweets. The mental health concerns on Twitter were geographically different in the UK. Among Twitter users who had mental health concerns during the COVID-19 pandemic, the male and age 21-60 group was dominant.

Before the COVID-19 pandemic, compared to rural people, British urban people had higher rates of CIS-R morbidity, a measure of mental disorder and depression ^16^. Moreover, the population density was positively correlated with the Twitter activity ^17^. These studies coincide with our geographic analysis results that the Great London metropolitan area, with the highest population and population density within the United Kingdom, has the highest proportion of Twitter users with mental health concerns, followed by Birmingham and Manchester metropolitan areas. All three major metropolitan areas have a higher percentage of Twitter users than the average level of the whole of England. In addition, Scotland appears to have a higher proportion of Twitter users with mental health concerns than the other three countries in the UK.

Similar to previous studies conducted on the general COVID-19-related tweets ^18 19^, our topic modeling results showed that the concern about the pandemic and dissatisfaction with government responses are the two major topics discussed in the mental health-related tweets, which also conforms with our conclusion from temporal analysis results. According to a recent study on tracking mental health and other symptoms mentioned on Twitter in the United States, monitoring discussions on Twitter related to mental health and other symptoms associated with the COVID-19 pandemic could provide a reliable indicator for the well-being during the public health crisis ^10^. Using tweets from the UK, we observed that the severity of the pandemic and the government’s strict measures to control the spread of the coronavirus could both affect the mental health concerns of British people—the latter’s impact might be more significant. In addition, by examining the representative tweets for each topic, we found that the COVID-19 skeptics, which argued that COVID-19 cases are hyped by the government or media to spread fear, took a non-negligible part in the mental health-related discussions. Moreover, the topics regarding the promotion of mental health care and support and relief for the vulnerable populations are also identified.

Our demographic inference results revealed an unnoticeable discrepancy in gender distribution on UK Twitter users who had mental health concerns. Among Twitter users who had mental health concerns, 56.9% were male, and 43.1% were female, which is almost identical to the estimated split of all UK Twitter users in 2017 (57% male and 43% female) ^20^. Therefore, the COVID-19 pandemic has a similar mental health impact between males and females. A previous study showed British Twitter users significantly skewed to the young people ^21^. Our study showed that the middle-aged and older adults, over the age of 30, roughly defined, take up 72% of Twitter users who had mental health concerns, while this group takes only 9.2% of all Twitter users in the United Kingdom ^21^. Therefore, it seems that the middle-aged and older adults in the UK were disproportionally more likely to have mental health concerns or express their mental health concerns through Twitter during the pandemic, which might result in their more vulnerability to the COVID-19 pandemic than the teenagers.

In this study, Twitter users who expressed mental health concerns did not necessarily have mental health issues. Additionally, only a small subset of Twitter users provided the geolocation information, as well as a valid human profile picture. Moreover, the user profile pictures may not come from their own. Demographic inference of Twitter users using deep learning algorithms may not be accurate. Therefore, these might lead to some biases in our results. In this study, we only focus on Twitter data and Twitter users, which may not necessarily represent the whole population in the UK.

## Conclusion and implications

Using Twitter data from the UK, we showed that there is a potential impact of the COVID-19 pandemic on mental health concerns, which varied among Twitter users with different geolocations and demographics. We have successfully demonstrated the potential of using social media to monitor and study mental health concerns during the COVID-19 pandemic, which is vital to protect public health, especially mental health.

## Data Availability

The Twitter data is publicly available and accessible to anyone with internet access.

## Acknowledgements

This study was supported by the University of Rochester CTSA award number UL1 TR002001 from the National Center for Advancing Translational Sciences of the National Institutes of Health. The content is solely the responsibility of the authors and does not necessarily represent the official views of the National Institutes of Health.

---

**Supplemental Figure 1.**
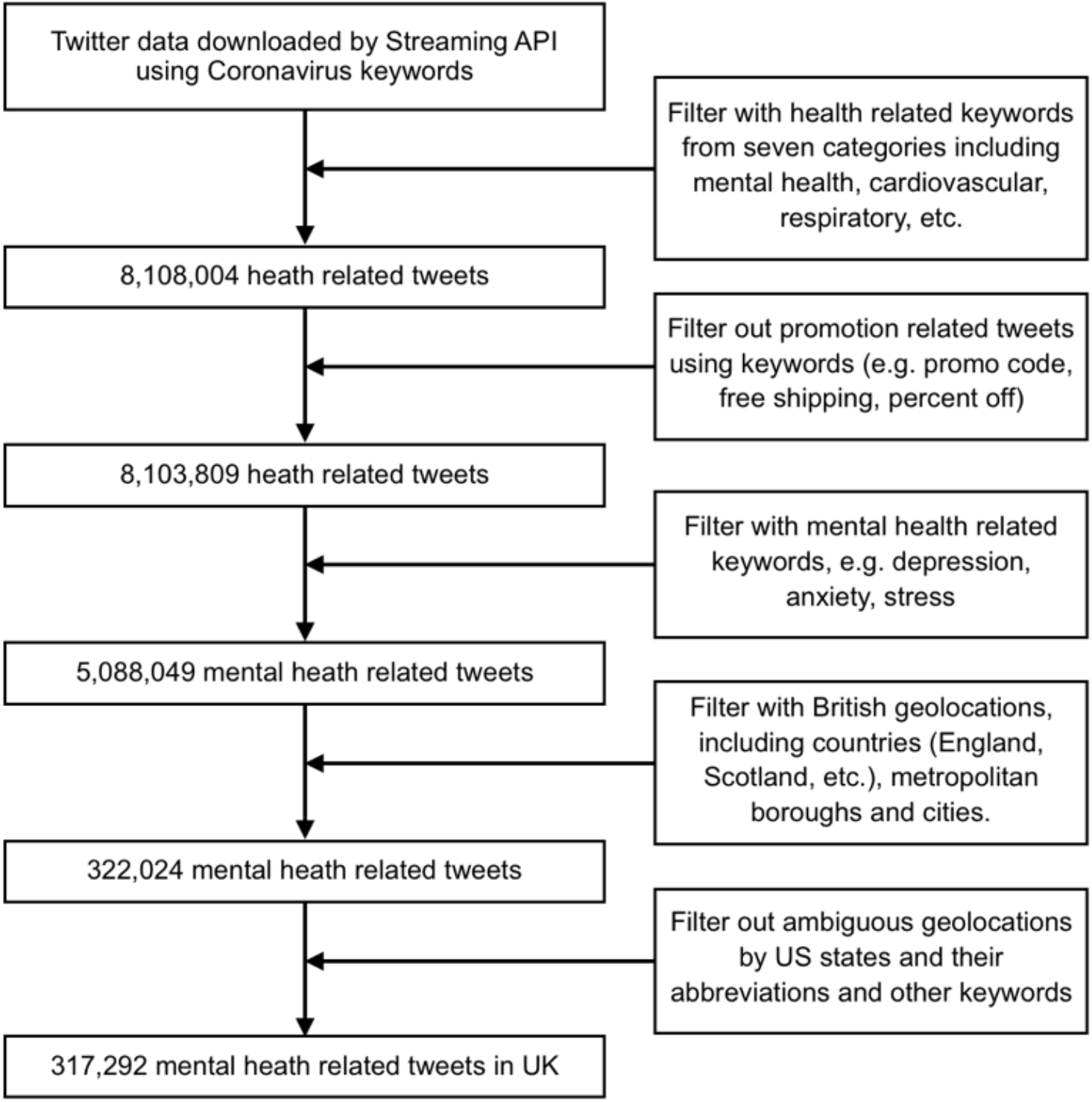
Flow chart of Twitter data pre-processing

**Supplemental Table 1.**
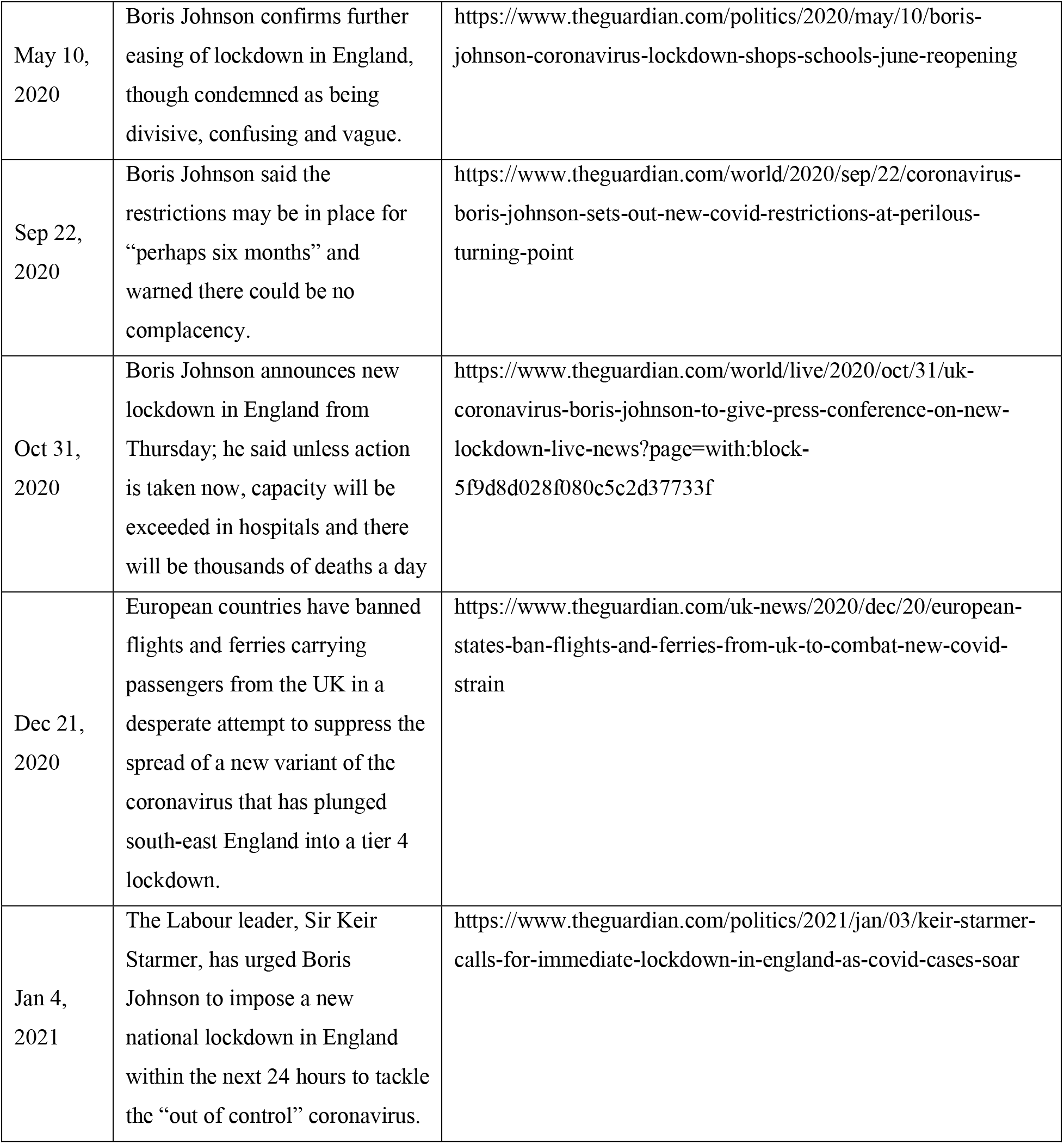
News associated with five peaks in mental-health-related tweets.

